# Circulating SFRP5 levels are elevated in colorectal cancer and correlate with overall survival in stage II-III disease

**DOI:** 10.1101/2024.03.14.24304260

**Authors:** Runhao Li, Saifei Liu, Kenny Yeo, Suzanne Edwards, Man Ying Li, Ryan Santos, Sima Kianpour Rad, Fangmeinuo Wu, Guy Maddern, Joanne Young, Yoko Tomita, Amanda Townsend, Kevin Fenix, Ehud Hauben, Timothy Price, Eric Smith

## Abstract

Secreted Frizzled-Related Protein 5 (SFRP5) modulates Wnt signalling pathways, affecting diverse biological processes. We assessed the diagnostic and prognostic value of circulating SFRP5 (cSFRP5) in colorectal cancer (CRC). Plasma cSFRP5 concentrations were measured using ELISA in healthy donors (n=133), individuals diagnosed with CRC (n=449), colorectal polyps (n=85), and medical conditions in other organs including cancer, inflammation, and benign states (n=64). Patients with CRC, polyps, and other conditions showed higher cSFRP5 levels than healthy individuals (p<0.0001). Receiver operating characteristic curves comparing healthy donors with medical conditions, polyps, and CRC were 0.814 (p<0.0001), 0.763 (p<0.0001), and 0.762 (p<0.0001), respectively. In CRC, cSFRP5 correlated with patient age (p<0.0001), tumour stage (p<0.0001), and histological differentiation (p=0.0273). Levels peaked in stage II versus I (p<0.0001), III (p=0.0007), or IV (p<0.0001), and were higher in stage III versus I (p=0.0007) and IV (p=0.0054), with no difference between I and IV. Elevated cSFRP5 levels predicted longer overall survival in stage II-III CRC (univariate: HR 1.82, 95% CI 1.02-3.26, p=0.024; multivariable: HR 2.34, 95% CI 1.12-4.88, p=0.015). This study confirms elevated cSFRP5 levels in CRC and reveals a correlation between elevated cSFRP5 and overall survival in stage II-III disease.

**Novelty and Impact:** Our study unveils the clinical significance of circulating Secreted Frizzled-Related Protein 5 (cSFRP5) as a potential diagnostic and prognostic marker in colorectal cancer (CRC). Analysing plasma from 731 participants, we provide novel insights into cSFRP5, observing elevated levels in CRC compared to healthy controls, with the highest concentrations in stage II. Importantly, elevated cSFRP5 levels correlate with prolonged survival in stage II-III patients, suggesting a promising avenue for improving outcomes and reducing recurrence.

## INTRODUCTION

Colorectal cancer (CRC) represents a significant global healthcare challenge, ranking as the third most diagnosed cancer and a leading cause of cancer-related mortality ^1^. With over 1.8 million reported cases and more than 0.9 million deaths in 2020 alone, there is urgency for the development of early detection strategies and to significantly improve treatment efficacy and survival rates. Enhancing screening methodologies, advocating regular screenings among high-risk populations, and developing precise, non-invasive screening technologies are imperative ^2,3^.

Secreted frizzled-related protein 5 (SFRP5) belongs to the SFRP family, an evolutionarily conserved group of extracellular glycoproteins crucial for modulating Wnt signalling pathways ^4^. These pathways play pivotal roles in various cellular processes, including embryonic development ^5^, tissue homeostasis, and the modulation of tumour initiation, growth, and metastasis ^6,7^. Structurally, SFRPs contain an amino-terminal signal peptide, a cysteine-rich domain (CRD), and a carboxy-terminal netrin-like domain (NTR) ^8^. The CRD is similar to the Wnt ligand binding site found in Frizzled family of cell surface receptor proteins ^9^. The NTR domain is involved in protein-protein interactions and is thought to contribute to the function of SFRPs ^10^. This structure enables SFRPs to act as soluble antagonists of Wnt signalling by interacting with Wnt ligands and inhibiting activation of the Frizzled receptors (reviewed in ^11^).

SFRP5, specifically, has been shown inhibit multiple Wnt ligands such as Wnt5A and Wnt11, thereby regulating both canonical and non-canonical Wnt pathways ^12,13^. Experimental evidence from murine disease models and studies on SFRP5 knockout mice suggests that secretion of SFRP5 by adipocytes prevents hepatic steatosis and metabolic dysfunction associated with liver fibrosis by modulating inflammatory cells within adipose tissue ^12,14^. In humans, downregulation of SFRP5 contributes to proinflammatory conditions in visceral adipose tissue, potentially exacerbating obesity-related comorbidities ^15^.

Indeed, circulating SFRP5 (cSFRP5) has emerged as a potential biomarker in various diseases, participating in diverse pathological processes ^16–18^. Typically, individuals with metabolic disorders like obesity, insulin resistance, and diabetes exhibit reduced levels of cSFRP5 compared to healthy controls ^16–19^. Meanwhile, interventions leading to significant weight loss in overweight individuals are associated with increased cSFRP5 levels, further highlighting its role in metabolic regulation ^20–22^. These findings collectively suggest that higher cSFRP5 levels are associated with reduced risk of metabolic disorders in humans. Interestingly, metabolic syndrome has been shown to be associated with increased risk of liver metastasis in CRC patients ^23^. Additionally, cSFRP5 has implications in cardiovascular diseases, with lower concentrations observed in individuals with arterial stiffness ^24^, atherosclerosis ^25^, coronary artery disease ^26,27^, and hypertension ^21,28^.

In contrast, the role of cSFRP5 in cancer remains understudied. Our previous study indicated that cSFRP5 was elevated in CRC compared to non-CRC controls, and elevated cSFRP5 levels were associated with extended disease-free survival among patients in stages I-III ^29^. Yet, limitations, such as small sample size and insufficient control groups, hindered the comprehensive understanding of its potential. In this study, we measured cSFRP5 concentrations in an independent, retrospective, multi-centre cohort encompassing 133 healthy controls without documented pathology, 449 patients with different stages of CRC, 85 individuals with colorectal polyps, and 64 individuals with other notable medical conditions in other organs including cancer, inflammation, and benign states. Our primary objectives were to validate cSFRP5’s diagnostic and prognostic potential and explore its correlation with CRC progression-related clinical-pathological parameters and overall survival. Our secondary objectives were to determine if cSFRP5 was altered in patients diagnosed with colorectal polyps and other medical conditions in other organs including cancer, inflammation, and benign states.

## MATERIALS AND METHODS

### Patient plasma samples

All plasma samples were obtained from Victorian Cancer Biobank (Table 1) and were sourced from multiple centres (Supplementary Table 1). The CRC samples were collected over a period from 1999 until 2021. All individuals had fasted from the previous night until after blood collection.

**Table 1.** Summary of participants in this study.

| **Group** | **n** | **Female (%)** | **Median age, years (range)** |
| --- | --- | --- | --- |
| Healthy (H) | 133 | 86 (64.7%) | 42 (16-80) |
| Notable medical conditions (D) | 64 | 15 (23.4%) | 60 (25-88) |
| Colorectal polyps (P) | 85 | 40 (47.1%) | 66 (22-88) |
| Colorectal Cancer (CRC) |  |  |  |
| Stage I | 147 | 60 (40.8%) | 68 (34-91) |
| Stage II | 103 | 40 (38.8%) | 68 (32-95) |
| Stage III | 109 | 36 (33.0%) | 68 (28-89) |
| Stage IV | 90 | 36 (40.0%) | 57.5 (25-82) |
| *CRC Total* | *449* | *172 (38.3%)* | *66 (25-95)* |
| **Total** | **731** | **313 (42.8%)** | **63 (16-95)** |

### SFRP5 enzyme-linked immunosorbent assay

The plasma cSFRP5 concentration was determined using a commercially sourced, commonly used enzyme-linked immunosorbent assay (ELISA) according to the manufacturer’s instructions (Cloud Clone Corp., TX, USA) ^29^. All ELISA kits used in this study were sourced from a single lot number (Lot# L210917454), with a detection range of 1.56-100 ng/mL. Plasma samples were diluted 1 in 2 in phosphate buffered saline (pH 7.0-7.2) and analysed in duplicate. The optical density at 450 nm was measured using iMark Microplate Absorbance Reader (Bio-Rad Laboratories, Inc., CA, USA), and the calculated cSFRP5 concentration was derived from the standard curve.

### Statistical analysis

Statistical analyses were conducted using either Prism 10 for MacOS (GraphPad Software, Inc., CA, USA), SAS On Demand for Academics (SAS Institute Inc., NC, USA), or the R package, survival v3.5. To assess differences in cSFRP5 concentration between patient groups, we utilised Kruskal-Wallis with Dunn’s multiple comparisons test. Correlations between cSFRP5 concentration and various prognostic clinical-pathological parameters were examined using the Mann-Whitney test. A linear regression model was employed to evaluate the relationship between cSFRP5 concentration and patient group, tumour invasion stage, TNM group, as well as other prognostic clinical-pathological parameters. This model was adjusted for potential confounding variables, including age and sex. Assumptions crucial to the linear models, such as normality of residuals and homoscedasticity, were found to be upheld through careful inspection of residual histograms and scatterplots of residuals against predicted values. Univariate and multivariable Cox proportional hazard models were performed using R package, survival v3.5, to assess relationship between overall survival, cSFRP5 concentration, TNM stage, age, sex, vascular or perineural invasion, histological differentiation, and tumour site. A p≤0.05 was deemed statistically significant.

## RESULTS

### Patient cohorts

The study included 731 participants (Table 1), comprising 133 healthy donors without documented pathology (Supplementary Table 2), 449 individuals diagnosed with CRC, 85 individuals with colorectal polyps (Supplementary Table 3), and 64 individuals with other notable medical conditions in other organs (Supplementary Table 4). The polyps group comprised the following subtypes: villous/tubulovillus (n=48), adenoma/adenomatous (n=31), sessile serrated (n=2), hyperplastic (n=2), inflammatory (n=1), and Peutz Jegher syndrome (n=1). The other notable medical conditions included patients diagnosed with cancer (n=4), chronic inflammation (n=49), and benign diseases (n=11). The healthy participants consisted of more females (p<0.0001) and were significantly younger (p<0.0001) (Supplementary fig. 1).

### Circulating SFRP5 was elevated in colorectal cancer

The cSFRP5 concentration was significantly higher in CRC (mean 17.67 ng/mL, 95% confidence interval (95% CI): 16.63-18.72 ng/mL) compared to healthy donors (9.26 ng/mL, 95% CI: 8.08-10.45 ng/mL; p<0.0001; Figure 1A). The area under the receiver operating characteristic (AUROC) curve for discriminating between CRC and healthy donors was 0.762 (95% CI: 0.717 to 0.807; p<0.0001; Figure 1B). With regards to TNM stage, the cSFRP5 was significantly higher in all stages of CRC compared to healthy donors (Figure 1C). Notably, the cSFRP5 levels were highest in stage II compared to either stage I (p<0.0001), III (p=0.0003), or IV (p<0.0001). The cSFRP5 was significantly elevated in stage III compared to stage I (p=0.0007) and IV (p=0.0054). There was no significant difference between stage I and IV. Recognising the notable variations in both age and sex among our patient cohorts and considering prior studies indicating the potential impact of these factors on cSFRP5 levels ^30,31^, we utilised a linear regression model to account for age and sex differences (Supplementary Table 5). Our analysis revealed a correlation between age and increased cSFRP5 concentration, but no discernible disparity was observed between males and females. Adjusting for age and sex did not alter the pattern of significant differences.

**Figure 1:**
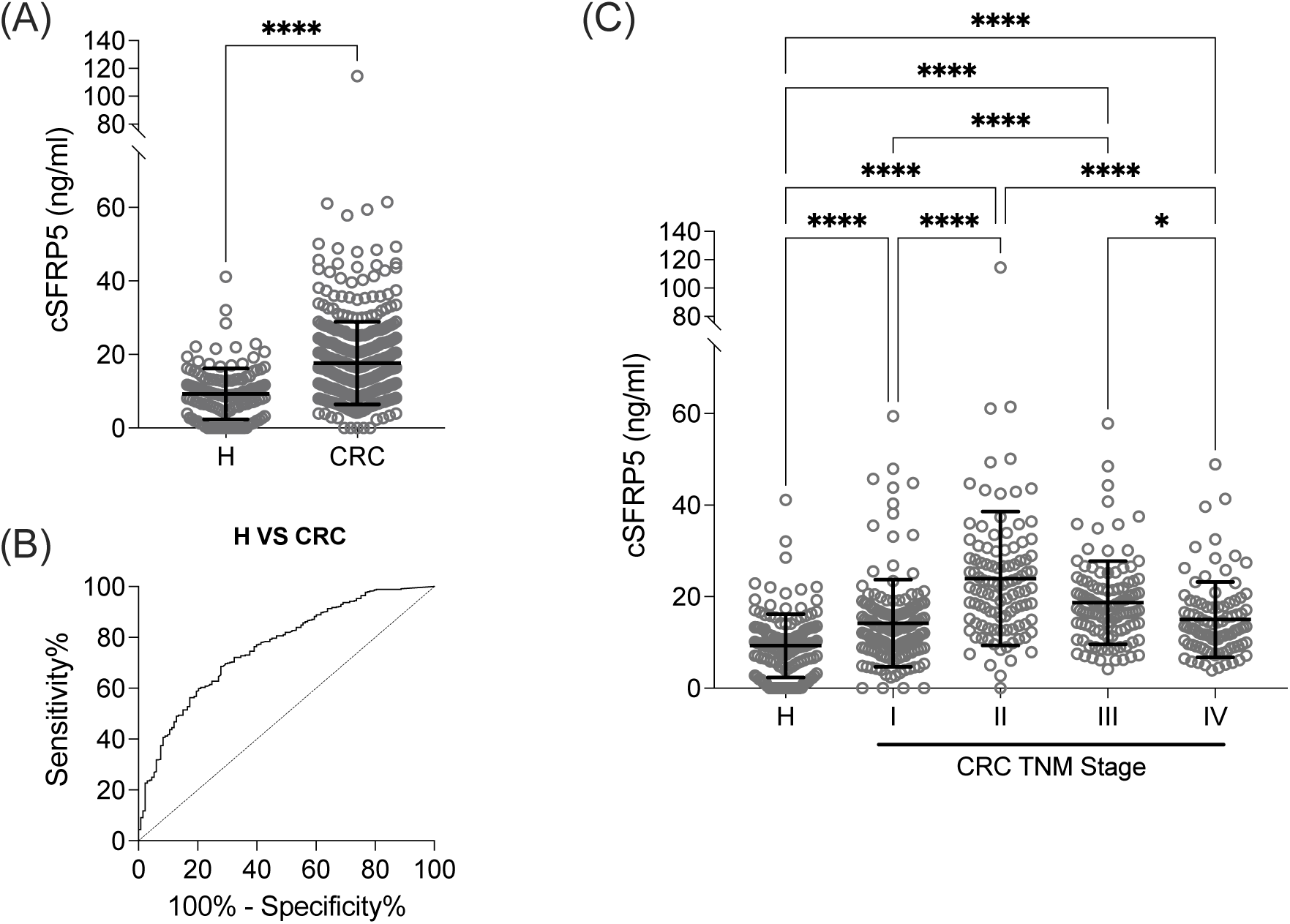
(**A**) Concentration of cSFRP5 in healthy donors (H) (n=133) and patients with colorectal cancer (CRC) (n=449). (**B**) AUROC curve for cSRFP5 concentration comparing H to CRC. (**C**) Concentration of cSFRP5 in healthy donors (H) (n=133) and CRC patients by TNM stage I (n=147), stage II (n=103), stage III (n=109), and stage IV (n=90). The data are the mean ± SD. Kruskal-Wallis with Dunn’s multiple comparisons were used, ****p<0.0001, *p<0.05.

### Circulating SFRP5 was elevated in colorectal polyps

Elevation of cSFRP5 was also observed in colorectal polyps, with a mean concentration of 17.34 ng/mL (95% CI: 15.00-19.69 ng/mL) compared to healthy donors (Supplementary fig. 2A; p<0.0001). The AUROC curve for discriminating polyps from healthy donors was 0.763 (95% CI: 0.699 to 0.828; p<0.0001; Supplementary fig. 2B). Furthermore, individuals with villous/tubulovillous (p<0.0001) and adenoma/adenomatous (p<0.01) polyps, categorized as having high cancer progression risk, displayed significantly elevated cSFRP5 levels compared to healthy controls (Supplementary fig. 2C).

### Circulating SFRP5 was elevated in other disease states

Comparing individuals with various medical conditions to healthy donors revealed elevated cSFRP5 levels among those with notable medical conditions, with a mean concentration of 21.23 ng/mL (95% CI: 17.84-24.62 ng/mL), significantly higher than healthy donors (p<0.0001; Supplementary fig. 3A). The AUROC curve for distinguishing between individuals with notable medical conditions and healthy donors yielded a value of 0.814 (95% CI: 0.746 to 0.882; p<0.0001; Supplementary fig. 3B). Moreover, cSFRP5 levels exhibited significant increases in individuals with other cancers (p<0.0001), inflammation (p<0.0001), and miscellaneous benign conditions (p<0.01) in other organs compared to healthy controls (Supplementary fig. 3C). However, it is important to acknowledge that the sample size of these subgroups was limited, necessitating further validation with larger cohorts.

### Elevated circulating SFRP5 levels were not exclusive to colorectal cancer

We employed a linear regression model to adjust for age and sex differences when comparing patient groups (Supplementary Table 6). The adjusted cSFRP5 concentration was notably higher in patients with significant medical conditions (p<0.0001), colorectal polyps (p=0.0020), and CRC (p<0.0001). Particularly, cSFRP5 levels were significantly elevated in patients with notable medical conditions compared to those with either colorectal polyps (p=0.0130) or CRC (p=0.0027). However, no significant difference was observed between the polyps and CRC groups. The AUROC curve for distinguishing between notable medical conditions and either CRC or polyps was 0.596 (95% CI: 0.518 to 0.674; p=0.0127) and 0.613 (95% CI: 0.519 to 0.706; p=0.0188), respectively (Supplementary Fig. 4). In summary, these findings suggest elevated cSFRP5 levels across various disease states, including colorectal polyps and CRC.

### Elevated circulating SFRP5 in colorectal cancer was associated with patient age, tumour stage and histological differentiation

In the patients with CRC, we investigated the correlations between the cSFRP5 concentration and different clinical-pathological parameters (Table 2 and Supplementary fig. 5). We observed a significant increase in cSFRP5 concentration with patient age (p<0.0001), TNM stage (p<0.0001), advanced depth of invasion (p<0.0001), in the absence of distant metastasis (p=0.0049), and with poorly differentiated tumours (p=0.0273). The cSFRP5 concentration was significantly higher in CRC patients with more invasive T3 and T4 primary tumours compared to the less invasive T1 and T2 (Supplementary fig. 5A). Although there was an increase in cSFRP5 concentration in CRC with mismatch repair deficiency (dMMR; Supplementary fig. 5I; p=0.0655), it did not reach statistical significance. We did not detect variations in relation to sex, lymph node metastasis, vascular or perineural invasion, diabetes, or hypertension in the patients with CRC.

**Table 2.** cSFRP5 concentration in CRC with various prognostic clinical-pathological parameters.

| Clinical parameters | n | Mean cSFRP5 (95% CI) | p value |
| --- | --- | --- | --- |
| **Age\*** | 449 |  | **<0.0001** |
| **Sex** |  |  |  |
| Female | 172 | 17.65 (15.97-19.33) | 0.3927 |
| Male | 277 | 17.69 (16.35-19.03) |  |
| **TNM stage** |  |  |  |
| I | 147 | 14.18 (12.63-15.74) | **<0.0001** |
| II | 103 | 23.94 (21.08-26.80) |  |
| III | 109 | 28.68 (16.96-20.41) |  |
| IV | 90 | 14.98 (13.26-16.70) |  |
| **Tumour invasion (T-stage)** |  |  |  |
| T1 | 54 | 15.19 (12.11-18.27) | **<0.0001** |
| T2 | 103 | 13.72 (12.10-15.34) |  |
| T3 | 175 | 21.15 (19.26-23.03) |  |
| T4 | 58 | 19.15 (16.34-21.95) |  |
| **Lymph node metastasis (N-stage)** |  |  |  |
| N0 | 260 | 18.06 (16.50-19.62) | 0.2122 |
| N1/N2 | 137 | 17.99 (16.49-19.49) |  |
| NR | 52 |  |  |
| **Distant metastasis (M-stage)** |  |  |  |
| M0 | 359 | 18.35 (17.13-19.57) | **0.0049** |
| M1 | 90 | 14.98 (13.26-16.70) |  |
| **Vascular or perineural invasion** |  |  |  |
| Negative | 381 | 17.43 (16.28-18.57) | 0.0980 |
| Positive | 68 | 19.07 (16.56-21.58) |  |
| **Histological differentiation** |  |  |  |
| Low (Grade 1-2) | 374 | 17.06 (16.03-18.09) | **0.0273** |
| High (Grade 3-4) | 75 | 20.74 (17.22-24.27) |  |
| **Tumour site** |  |  |  |
| Colon | 311 | 18.07 (16.72-19.42) | 0.7549 |
| Rectum | 138 | 16.78 (15.25-18.31) |  |
| **Mismatch repair status** |  |  |  |
| pMMR | 220 | 18.21 (16.79-19.63) | 0.0655 |
| dMMR | 24 | 25.20 (15.80-34.60) |  |
| NR | 205 |  |  |
| **Diabetes** |  |  |  |
| Negative | 130 | 17.66 (15.40-19.91) | 0.2007 |
| Positive | 94 | 18.47 (16.43-20.50) |  |
| NR | 225 |  |  |
| **Hypertension** |  |  |  |
| Negative | 26 | 16.20 (13.93-18.46) | 0.8196 |
| Positive | 198 | 18.23 (16.50-19.96) |  |
| NR | 225 |  |  |

*\*Analysed as a continuous variable.*
*Mismatch repair deficiency (dMMR)*
*Mismatch repair proficient (pMMR)*
*Not recorded (NR)*
*95% confidence interval (95% CI)*

### Circulating SFRP5 correlates with overall survival in stage II-III colorectal cancer

We examined the relationship between cSFRP5 and overall survival of all CRC patients using Cox proportional hazard models (Supplementary Table 7). The median follow-up period was 4.3 years (range 0.08-20.7 years). Patients were stratified into low and high groups according to the median cSFRP5 concentration of 16.34 ng/mL observed among all CRC patients included in this study. When considering the entire CRC population encompassing stages I-IV, no significant association emerged between cSFRP5 concentration and overall survival, as revealed by both univariate (p=0.4866) and multivariable (p=0.9721) models (Supplementary Fig. 6 and Supplementary Table 7).

Ultimately, we examined the relationship between cSFRP5 and overall survival among stage II-III CRC patients, who are at an elevated risk of disease recurrence. High cSFRP5 levels were associated with extended overall survival (Figure 2 and Table 3; HR 1.82; 95% CI 1.02-3.26; p=0.0442). This correlation persisted in multivariable analysis, accounting for confounding factors (HR 2.34; 95% CI 1.12-4.88; p=0.0229). These findings underscore the prognostic relevance of cSFRP5 in CRC, particularly in the context of stage II-III disease.

**Figure 2:**
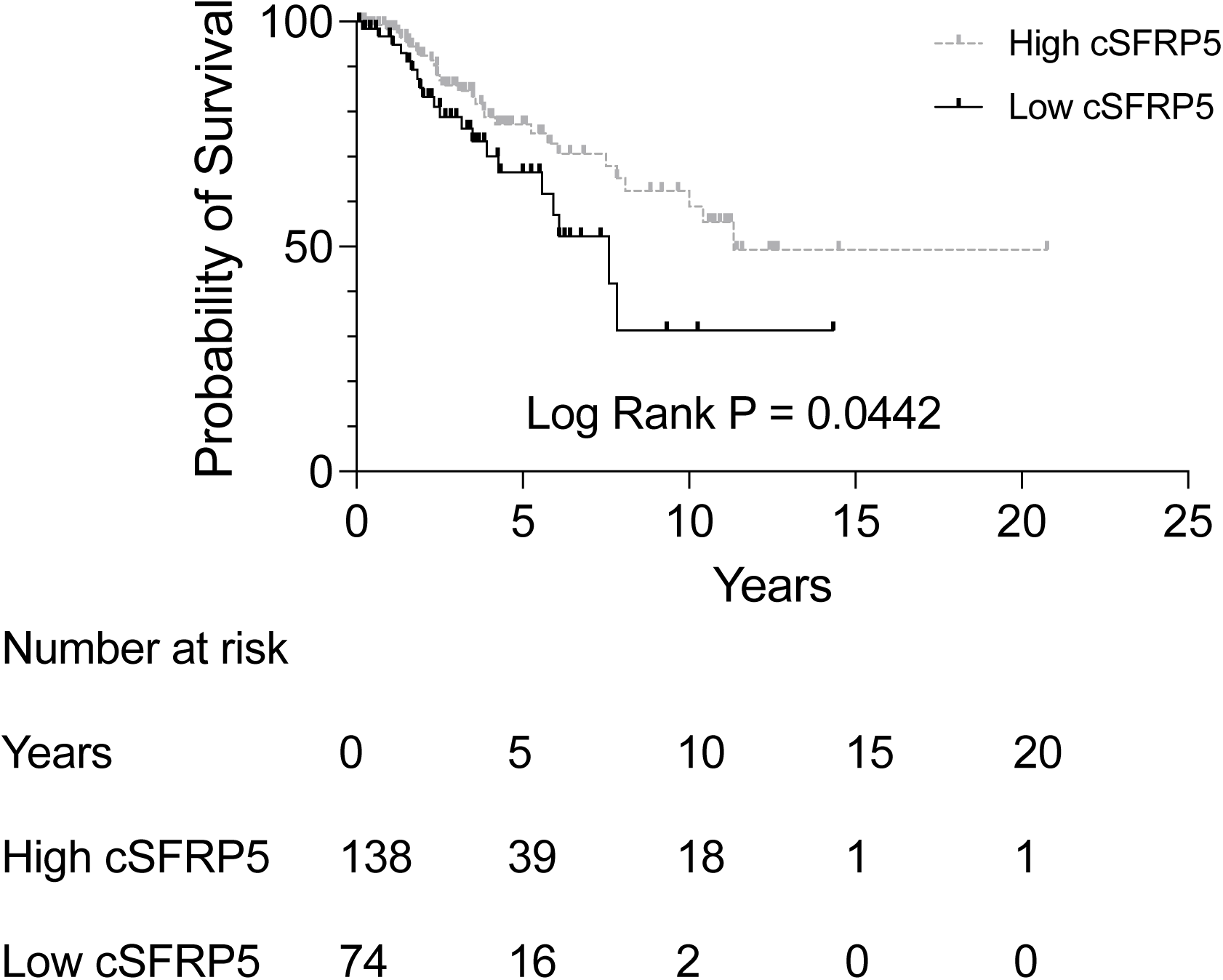
Kaplan-Meier survival curve for overall survival in patients with stage II-III colorectal CRC (n=212) dichotomized into low (≤16.34 ng/mL), or high (>16.34 ng/mL) cSFRP5 concentration.

**Table 3.** Univariate and multivariable analysis of overall survival in stage II-III CRC.

| Predictor | Univariate |  | Multivariable |  |
| --- | --- | --- | --- | --- |
|  | Hazard ratio (95% CI) | p value | Hazard ratio (95% CI) | p value |
| **cSFRP5** |  |  |  |  |
| High (>16.34 ng/mL) | 1 |  | 1 |  |
| Low (≤16.34 ng/mL) | 1.82 (1.02-3.26) | **0.0442** | 2.34 (1.12-4.88) | **0.0229** |
| **Age\*** | 1.06 (1.03-1.09) | **<0.001** | 1.07 (1.03-1.10) | **<0.001** |
| **Sex** |  |  |  |  |
| Male | 1 |  | 1 |  |
| Female | 0.80 (0.44-1.45) | 0.4609 | 0.81 (0.43-1.53) | 0.5210 |
| **TNM stage** |  |  |  |  |
| II | 1 |  | 1 |  |
| III | 1.41 (0.80-2.47) | 0.2366 | 1.18 (0.61-2.27) | 0.6151 |
| **Differentiation** |  |  |  |  |
| Low (Grade 1-2) | 1 |  | 1 |  |
| High (Grade 3-4) | 1.17 (0.63-2.16) | 0.6203 | 1.03 (0.53-1.92) | 0.9920 |
| **Vascular or perineural invasion** |  |  |  |  |
| Negative | 1 |  | 1 |  |
| Positive | 1.67 (0.89-3.11) | 0.1065 | 1.78 (0.89-3.55) | 0.1003 |
| **Tumour site** |  |  |  |  |
| Colon | 1 |  | 1 |  |
| Rectum | 0.79 (0.43-1.43) | 0.4344 | 0.95 (0.51-1.77) | 0.8839 |

\*Analysed as a continuous variable.

## DISCUSSION

This study investigated the levels of cSFRP5 across a diverse cohort, including healthy individuals, as well as those with CRC, colorectal polyps, and various other medical conditions. Elevated concentrations of cSFRP5 were identified in patients with CRC, polyps, and other significant illnesses, compared to the healthy controls. Notably, among CRC patients, cSFRP5 levels were highest in TNM stage II, followed by stage III, and were lowest in both stage I and IV. Furthermore, cSFRP5 levels exhibited an increase with patient age and were higher in the poorly differentiated tumours. Moreover, high cSFRP5 levels were associated with longer overall survival in the stage II-III CRC patients.

In our previous study, we highlighted the potential of cSFRP5 concentration as a diagnostic marker, effectively distinguishing CRC patients from those without CRC (AUROC 0.828, 95% CI 0.761-0.898) ^29^. In the current study we reported that the cSFRP5 level comparing patients with CRC to healthy donors had a AUROC of 0.762 (95% CI: 0.717 to 0.807; p<0.0001). However, it is important to emphasise that we did not find a significant difference between patients with colorectal polyps and those with CRC. Noteworthy is the fact that most of these individuals with polyps have a high risk of developing into cancer. This suggests that cSFRP5 may hold value in identifying individuals with high-risk polyps. However, our latest research revealed that individuals with other notable medical conditions also exhibited elevated cSFRP5 levels. These findings raise concerns about the exclusive use of cSFRP5 as a diagnostic biomarker for CRC population screening. However, it is important to note that the notable medical conditions group consisted of patients admitted to hospitals for treatment of various diseases, including cancer, inflammation, and benign states in organs other than the colorectum, and do not necessarily represent the general population.

Regarding CRC prognosis, our findings indicate that cSFRP5 levels are most elevated in patients with TNM stage II tumours, followed by stage III, and are at their lowest in both stage I and IV. This was similar to our previous findings in an independent CRC cohort, except that cSFRP5 levels were elevated in stage I, II and III compared to stage IV ^29^. It is important to note that the previous study was constrained by a smaller sample size. Additionally, we previously reported an association between elevated cSFRP5 and longer disease-free survival (HR 2.385; 95% CI 1.181–4.816; p=0.015) ^29^. Unfortunately, the current study lacked data on disease recurrence for a comprehensive analysis of disease-free survival. However, the current observation that elevated cSFRP5 in CRC patients is linked to improved overall survival is consistent with the previous findings for disease-free survival. Taken together, these findings suggest a favourable association between elevated cSFRP5 in CRC and survival outcomes, particularly in those with stage II-III disease.

The explanation for the increased cSFRP5 observed in patients with CRC, polyps, and in the other disease states remains largely unclear. In CRC, it is well established that SFRP5 transcript expression decreases due to promoter DNA hypermethylation ^29,32^, suggesting that the cancer cells are unlikely to be the origin of the heightened cSFRP5. However, to date there have been no studies that have reported SFRP5 protein levels in colorectal tissues. Research indicates that pancreatic cells express some of the highest levels of SFRP5 ^29,33,34^. Elevated glucose and insulin levels have been shown to decrease SFRP5 expression in pancreatic cells ^35^, and insulin infusion leads to a decrease in serum cSFRP5 ^36^. Conversely, free fatty acids counteract the insulin-induced decrease in cSFRP5 ^36^. Furthermore, serum cSFRP5 is positively correlated with HDL-cholesterol and negatively correlated with triglycerides and adiponectin ^36^. Additionally, cSFRP5 is inversely associated with circulating levels of pro-inflammatory cytokines, including IL-6 and TNFα ^37^. These collective observations suggest that systemic disruptions in metabolism and inflammation drive changes in cSFRP5 levels, and potentially account for the elevated concentrations observed in the study.

It is well accepted that SFRP5 binds Wnt ligands to antagonise Wnt signalling pathways ^4^. The Wnt signalling pathway is intricate and contingent on context. This pathway is indispensable for normal cellular processes, including cell proliferation, differentiation, and tissue homeostasis. However, dysregulation of the Wnt pathway is a pivotal factor in the development and progression of CRC. The non-canonical Wnt ligand, Wnt5a, is overexpressed in CRC and promotes cancer cell epithelial-mesenchymal transition, facilitating metastasis ^38,39^. The primary sources of Wnt5a in CRC are fibroblasts and macrophages. Hirashima et al. demonstrated that Wnt5a was expressed by cancer associated fibroblasts ^40^. The Wnt5a+ cancer associated fibroblasts were significantly associated with TNM stage, and recurrence. Subsequent *in vitro* analyses using human recombinant Wnt5a protein revealed that cancer cell proliferation and migration were significantly increased by stimulation with Wnt5a. Liu et al. identified that Wnt5a+ tumour-associated macrophages were associated with disease progression and poor prognosis ^41^. Moreover, Wnt5a+ macrophages promoted CRC cell proliferation, invasion, and migration, and the knockdown of Wnt5a significantly impaired the pro-tumour functions of tumour-associated macrophages. We propose that the observed elevated levels of cSFRP5 inhibit Wnt5a-mediated disease progression. However, we acknowledge that further studies are required.

We noticed higher cSFRP5 levels in mismatch repair deficient CRC compared to proficient ones, but it didn’t reach statistical significance. Unfortunately, only 244 out of 449 (54.3%) patients had their mismatch repair status reported, as it was not routinely tested at these institutions. Mismatch repair deficient CRC tends to have more mutations, generating neoantigens that attract immune cells to the tumour microenvironment. Meanwhile, SFRP5 by virtue of antagonising Wnt signalling pathways, has various effects on immune cells, influencing their behaviour and differentiation ^42^. It can shift macrophages toward an anti-inflammatory type ^12^ and affect the balance between effector and regulatory T cells, contributing to immune stability ^43^. Overall, SFRP5 appears to play a role in modulating immune cell differentiation, affecting the balance between pro-inflammatory and anti-inflammatory responses. More research is needed to determine the specific role of cSFRP5 in shaping the tumour immune cell microenvironment in CRC.

SFRP5 has garnered attention as a promising therapeutic target for various conditions, including metabolic diseases ^16^. Notably, SFRP5 administration reversed hyperglycaemia and hepatic steatosis in multiple mouse models of metabolic dysfunction ^12^. Lifestyle modifications leading to substantial weight loss in overweight individuals have been reported to increase cSFRP5 levels ^20–22^. Exercise training induced calorie expenditure in individuals with type 2 diabetes mellitus raised cSFRP5 levels ^44^. Additionally, clinically prescribed drugs such as the glucagon-like peptide-1 receptor agonist liraglutide have demonstrated the ability to elevate cSFRP5 levels ^31^. Considering the observed association of elevated cSFRP5 with improved disease-free survival ^29^ and overall survival, interventions that elevate cSFRP5 and target Wnt signalling pathways could emerge as attractive options for personalized medicine, particularly as an adjuvant therapy in individuals with stage II-III CRC with a heightened risk of disease recurrence.

A significant limitation of this study is its retrospective design, which precludes adjustments to clinical practices. Additionally, the absence of complete data for potentially important confounders, such as hypertension, obesity, insulin resistance, and diabetes represent a noteworthy drawback. The unavailability of anthropometric measurements, such as body weight, height, and waist circumference, further constrains the comprehensiveness of our analysis. Moreover, the lack of information on therapeutic drug usage, such as chemotherapeutics and anti-inflammatories, limits our ability to adequately assess these factors. Moving forward, the validation of our findings through prospective studies involving comprehensive CRC patient cohorts is crucial for advancing our understanding of the diagnostic and prognostic potential of cSFRP5.

In conclusion, this study validated previous findings that cSFRP5 levels are elevated in CRC patients and revealed that the highest cSFRP5 concentrations were detected in TNM stage II, followed by stage III, and the lowest in stages I and IV. Importantly, we report the novel finding that elevated cSFRP5 levels correlated with extended overall survival among stage II-III CRC patients, emphasizing its potential clinical significance as a prognostic biomarker. Our findings suggest that augmenting cSFRP5 levels could potentially enhance survival outcomes for stage II-III CRC patients, who are most vulnerable to disease recurrence.

## Data Availability

The data that support the findings of this study are available from the corresponding author upon reasonable request.

## ADDITIONAL INFORMATION

### Acknowledgements

The authors wish to thank the study participants, nurses and clinicians from Victorian Cancer Biobank in Melbourne, Victoria, Australia.

## Author Contributions

Conceptualization: Runhao Li, Timothy Price, Ehud Hauben and Eric Smith; Methodology: Runhao Li, Saifei Liu, Sima Kianpour Rad, Ryan Santos, Man Ying Li, Fangmeinuo Wu; Formal analysis and investigation: Runhao Li, Kenny Yeo, Suzanne Edwards, Eric Smith; Writing - original draft preparation: Runhao Li; Writing - review and editing: Runhao Li, Kenny Yeo, Suzanne Edwards, Ryan Santos, Amanda Townsend, Yoko Tomita, Guy Maddern, Joanne Young, Kevin Fenix, Ehud Hauben, Timothy Price, and Eric Smith; Funding acquisition: Timothy Price, Ehud Hauben, and Eric Smith; Resources: Timothy Price, Ehud Hauben, and Eric Smith; Supervision: Kevin Fenix, Timothy Price, and Eric Smith.

## Ethics approval and consent to participate

This study was performed in line with the principles of the Declaration of Helsinki, and in accordance with the Australian Code for the Responsible Conduct of Research and the National Statement on Ethical Conduct in Human Research. Approval was granted by the Central Adelaide Local Health Network Human Research Ethics Committee (Date 21-Sep-2021/No. HREC/14/TQEHLMH/164). Informed consent was obtained from all individual participants included in the study.

## Abbreviations

(AUROC): area under the receiver operating characteristic
(cSFRP5): circulating SFRP5
(CRC): colorectal cancer
(CRD): cysteine-rich domain
(ELISA): enzyme-linked immunosorbent assay
(dMMR): mismatch repair deficiency
(pMMR): mismatch repair proficient
(NTR): netrin-like domain
(SFRP5): secreted frizzled-related protein 5

---

**Supplementary fig. 1:**
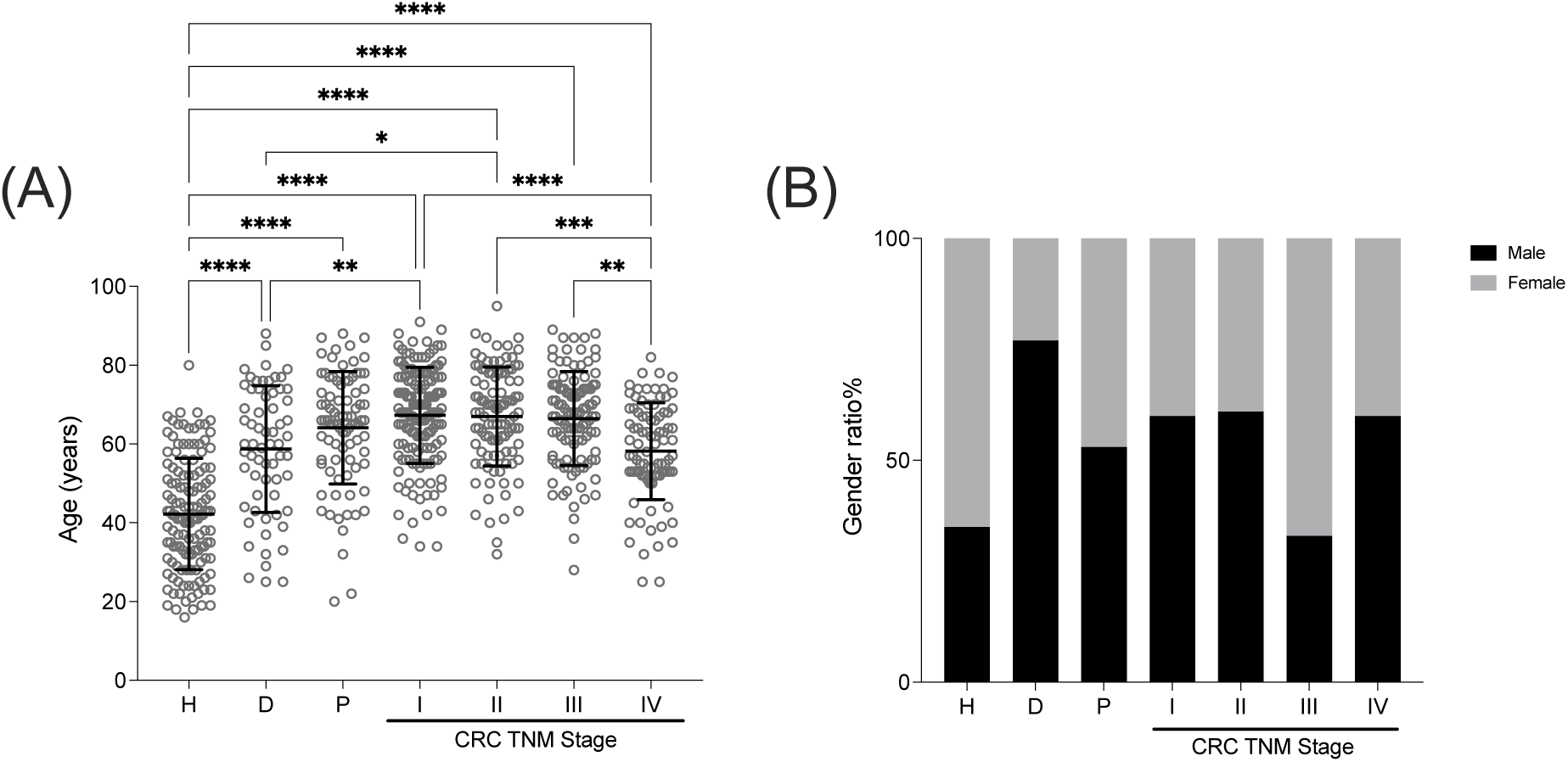
Age and gender of individuals in different groups. (**A**) Comparison of age in healthy donors without documented pathology (H) (n=133), patients with other notable medical conditions (D) (n=64), colorectal polyps (P) (n=85), or colorectal cancer (CRC) patients (n=449): stage I (n=147), stage II (n=103), stage III (n=109), and stage IV (n=90). Data as mean ± SD. Kruskal-Wallis with Dunn’s multiple comparisons were used, ****p<0.0001, ***p<0.001, **p<0.01, *p<0.05. (**B**) Comparison of gender ratio between different groups.

**Supplementary fig. 2:**
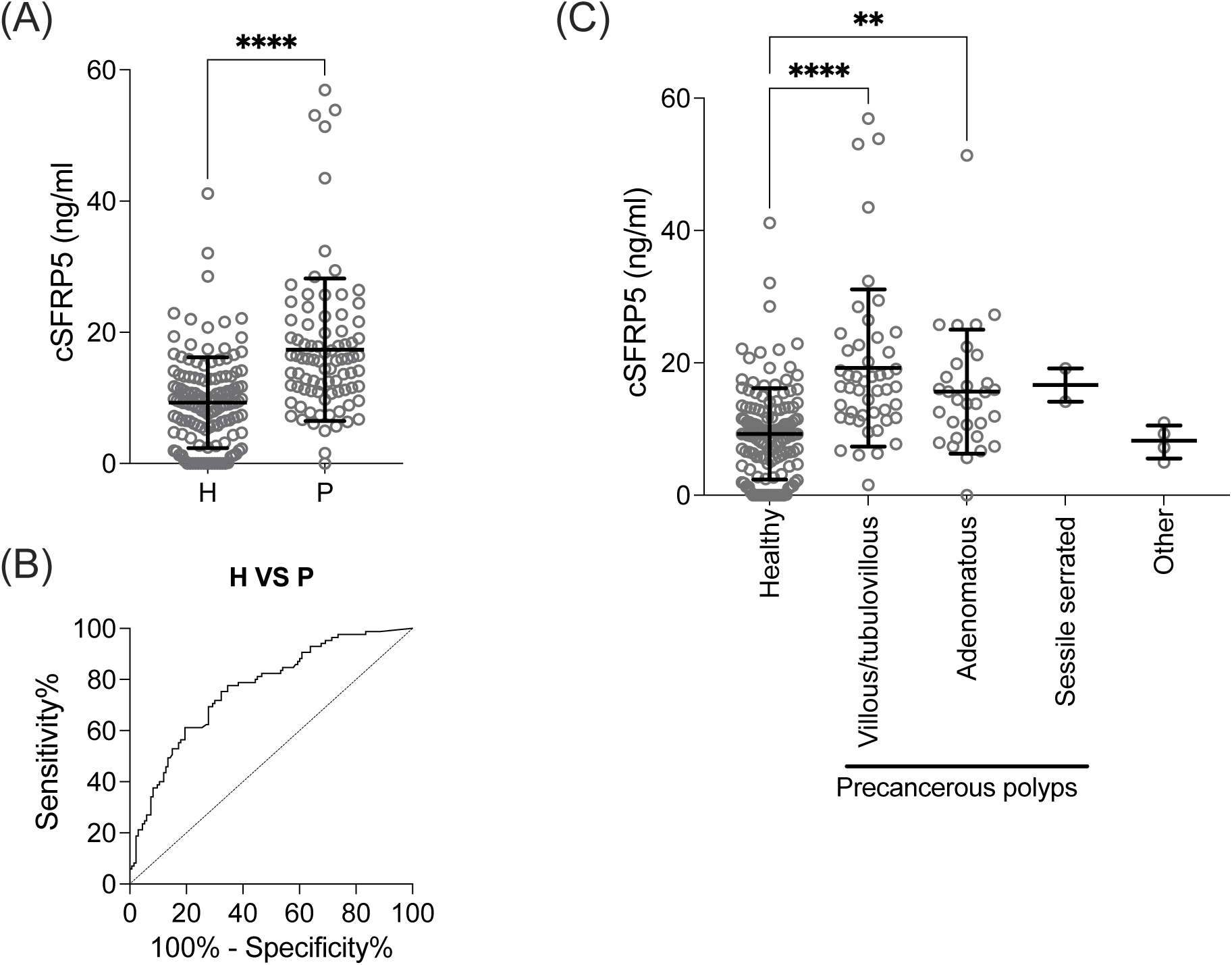
Concentration of cSFRP5 from healthy donors (H) and patients with colorectal polyps (P) (**A**) Comparison of cSFRP5 concentration in H (n=133) and P (n=85). (**B**) AUROC curve for cSRFP5 concentration comparing H to P. (**C**) Concentration of cSFRP5 in healthy donors (H) and patients with villous/tubulovillous (n=48), adenoma/adenomatous (n=31), sessile serrated (n=2), and other (n=4) types of colorectal polyps. The data are the mean ± SD. Kruskal-Wallis with Dunn’s multiple comparisons were used, ****p<0.0001, **p<0.01.

**Supplementary fig. 3:**
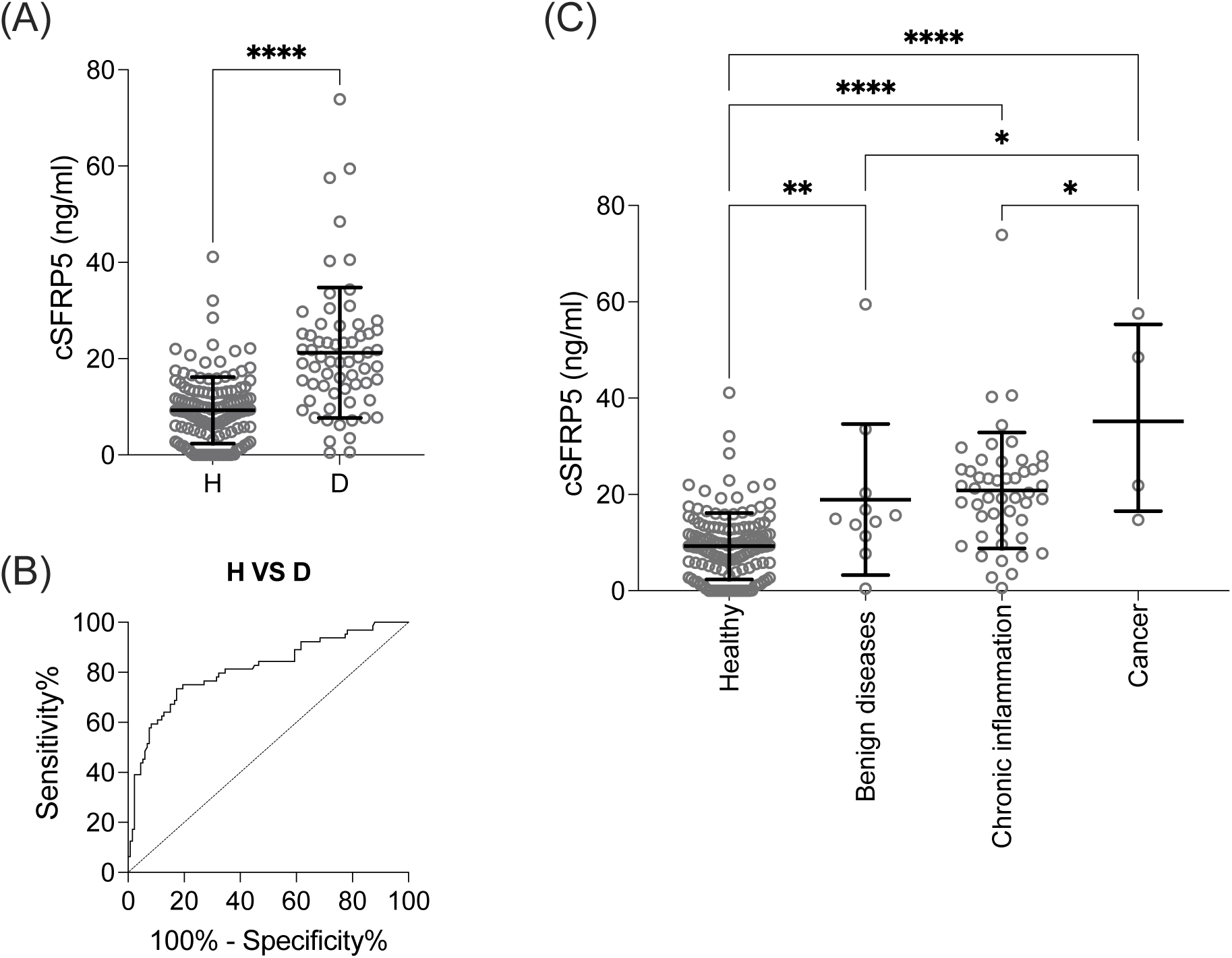
Concentration of SFRP5 from healthy donors (H) and other notable medical conditions (D). (**A**) Comparison of cSFRP5 concentration in H (n=133) and D (n=64). (**B**) AUROC curve for cSRFP5 concentration comparing H to D. (**C**) Concentration of cSFRP5 in healthy donors (H) (n=133) and patients with different types of notable medical conditions or significant disease by benign diseases (n=11), chronic inflammation (n=49), and cancer (n=4). The data are the mean ± SD. Kruskal-Wallis with Dunn’s multiple comparisons were used, ****p<0.0001, ***p<0.001, **p<0.01, *p<0.05.

**Supplementary fig. 4:**
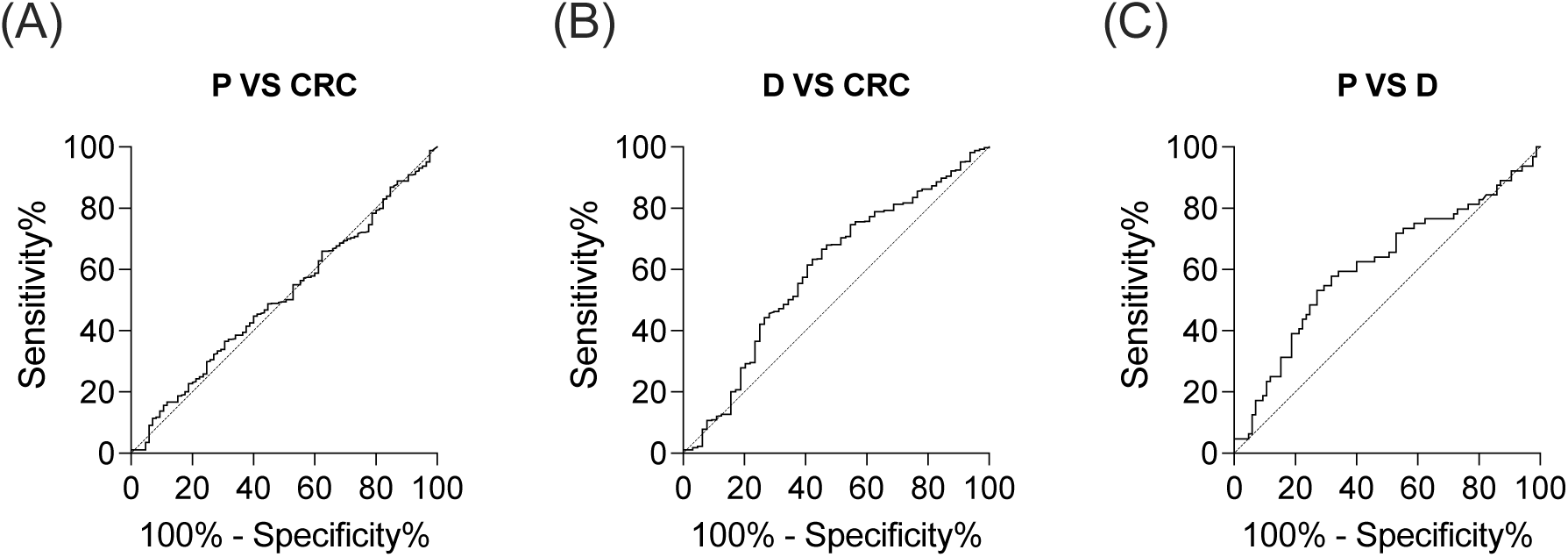
cSFRP5 AUROC curves for (**A**) colorectal cancer polyps (P) (n=85) versus colorectal cancer (CRC) (n=449), (**B**) other notable medical conditions (D) (n=64) versus CRC (n=449), and (**C**) P (n=85) versus D (n=64).

**Supplementary fig. 5:**
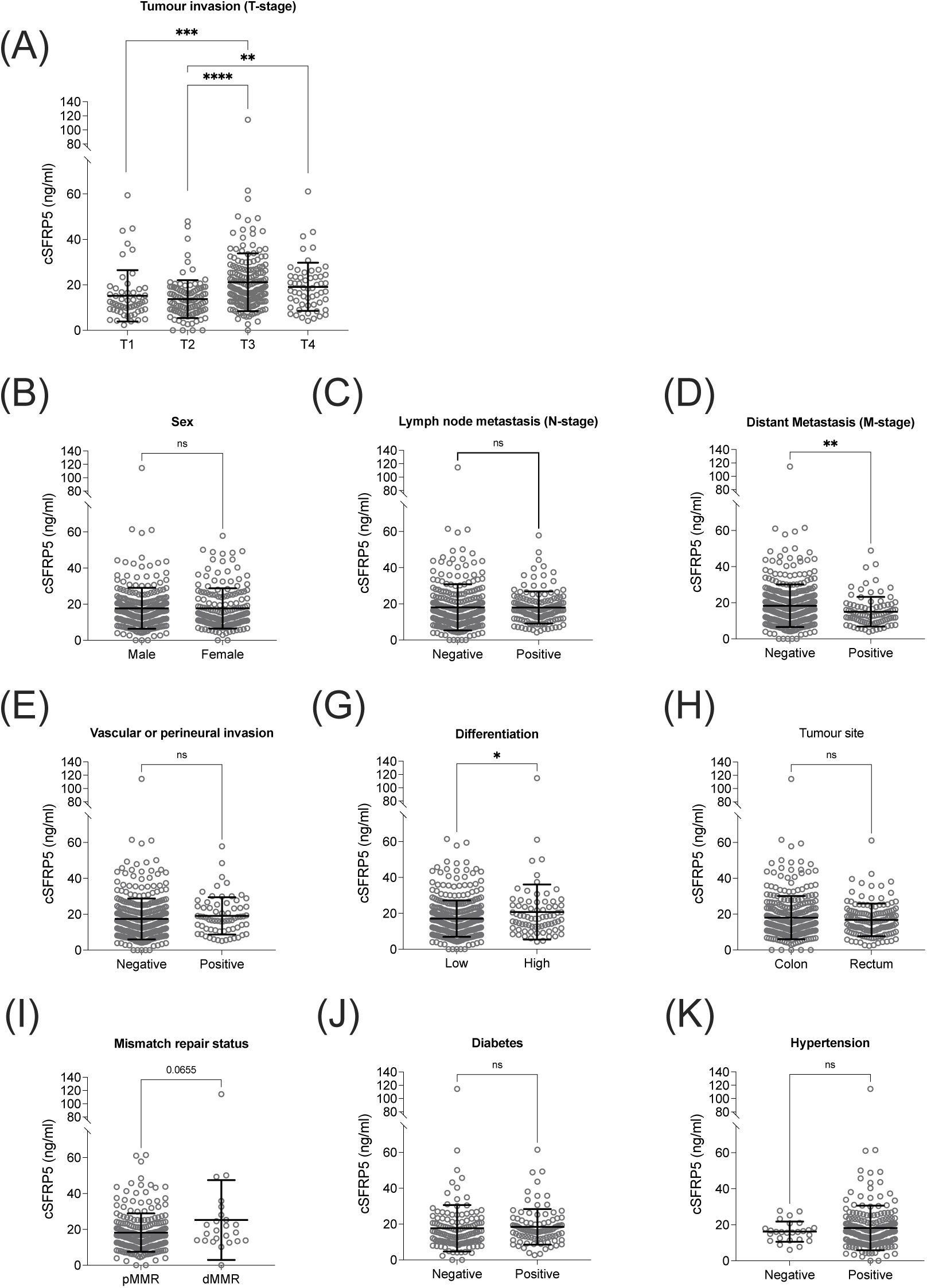
Concentration of cSFRP5 by various prognostic clinical-pathological parameters. (A) primary tumour invasion (T) stage. T1: tumour invades submucosa (n=54), T2: tumour invades muscularis propria (n=103), T3: tumour invades through the muscularis propria into peri-colorectal tissues (n=175), and T4: tumour directly invades or is adherent to other organs or structures (n=58). (B-K) cSRFP5 concentration comparison of sex, lymph node metastasis, distant metastasis, vascular or perineural invasion (VPNI), differentiation, tumour site, mismatch repair status, diabetes, and hypertension respectively. The data are the mean ± SD. A linear regression model was used for multiple comparisons, ****p<0.0001, ***p<0.001, **p<0.01, *p<0.05.

**Supplementary fig. 6:**
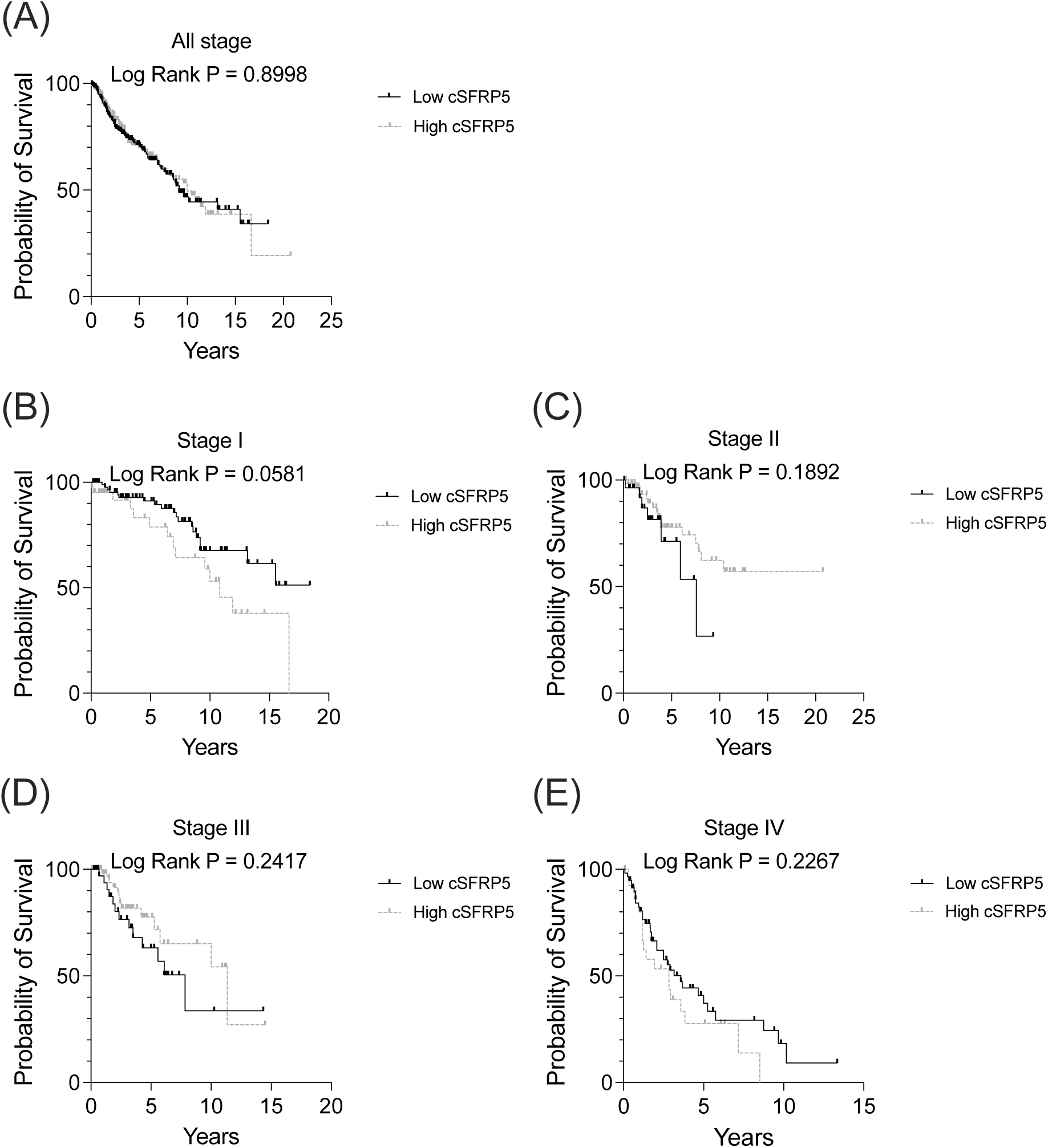
The Kaplan-Meier survival curves for overall survival in patients with colorectal cancer based on low and high cSFRP5 expression levels. (**A**) All patients (n=449), (**B**) Stage I (n=147), (**C**) Stage II (n=103), (**D**) Stage III (n=109), (**E**) Stage IV (n=90). Patients are dichotomized into low (≤16.34 ng/mL), or high (>16.34 ng/mL) cSFRP5 concentration, and survival curve were compared using Kaplan-Meier plots.

**Supplementary Table 1.** Summary of all samples from different institutions.

| **Hospital / Institution** | **Healthy**<br>**(H)** | **Notable**<br>**medical**<br>**conditions (D)** | **Colorectal**<br>**polyps (P)** | **Colorectal**<br>**Cancer (CRC)** | **Total (n)** |
| --- | --- | --- | --- | --- | --- |
| Austin Hospital (AH) | 21 | 26 | 8 | 85 | 140 |
| Barwon Health (BH) | - | - | 3 | 1 | 4 |
| Eastern Health (EH) | 6 | 11 | 20 | 184 | 221 |
| Monash Health (MH) | - | - | 4 | 5 | 9 |
| North Health (NH) | 2 | 6 | 5 | 3 | 16 |
| Peter MacCallum<br>Cancer Centre (PM) | 104 | 21 | 15 | 86 | 226 |
| Royal Melbourne<br>Hospital (RMH) | - | - | 13 | 20 | 33 |
| Western Health (WH) | - | - | 17 | 65 | 82 |
| **Total** | **133** | **64** | **85** | **449** | **731** |

**Supplementary Table 2.** Summary of healthy donors without significant documented pathology.

| **Group** | **n** | **Female (%)** | **Median age, years**<br>**(range)** |
| --- | --- | --- | --- |
| Healthy | 75 | 28 (37.3%) | 41 (16-80) |
| Prophylactic mastectomy | 37 | 37 (100%) | 41 (22-60) |
| Breast reduction | 19 | 19 (100%) | 43 (18-64) |
| Uterus endocervical polyp | 1 | 1 (100%) | 63 |
| Lipoma | 1 | 1 (100%) | 68 |
| **Total** | **133** | **86 (50.0%)** | **42 (16-80)** |

**Supplementary Table 3.** Summary of patients with colorectal polyps.

| **Polyp type** | **n** |
| --- | --- |
| Villous/tubulovillous adenoma | 48 |
| Adenoma/adenomatous | 31 |
| Sessile serrated | 2 |
| Hyperplastic | 2 |
| Inflammatory | 1 |
| Peutz Jegher syndrome | 1 |
| **Total** | **85** |

**Supplementary Table 4.**
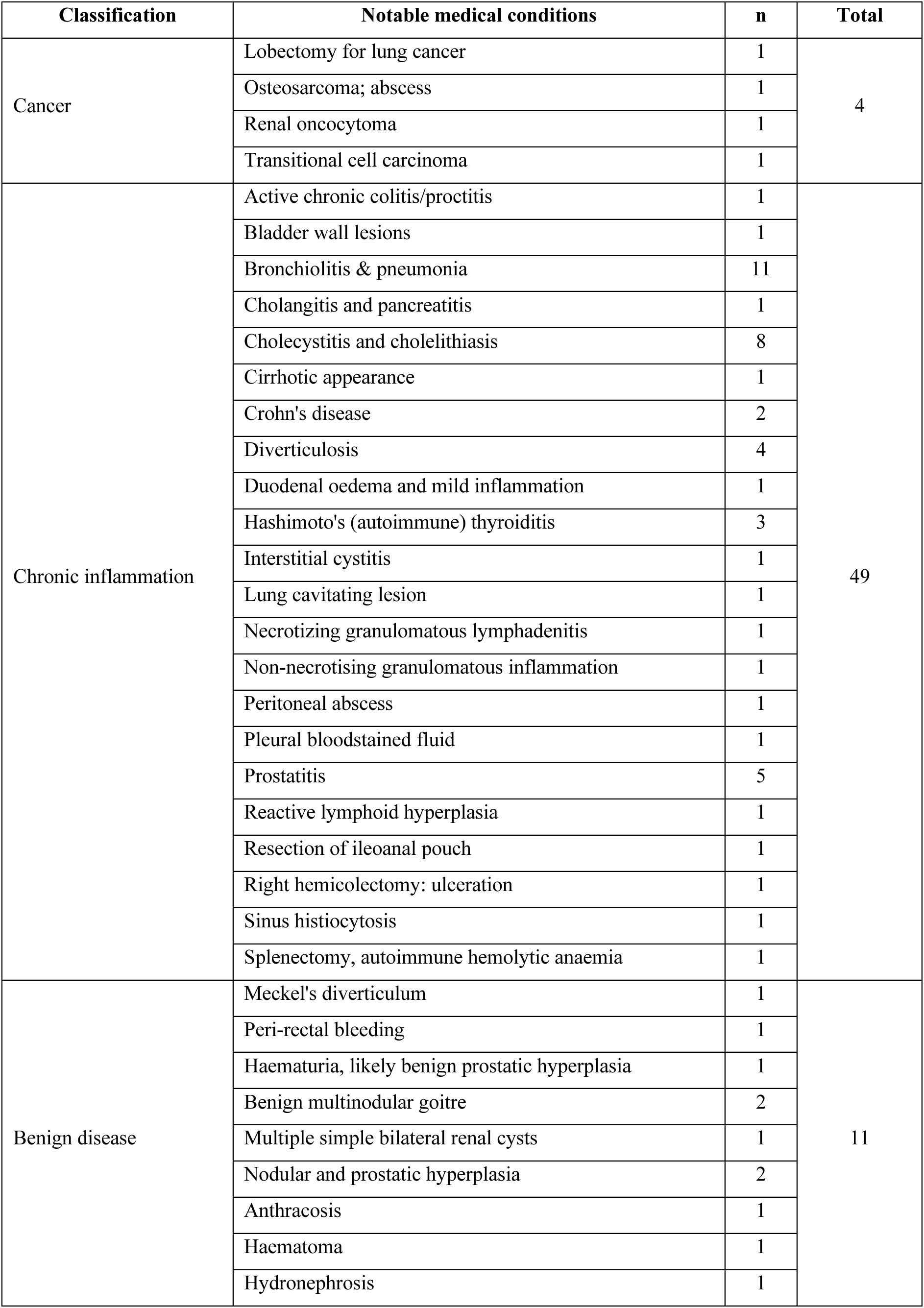
Summary of patients with notable medical conditions.

**Supplementary Table 5.** Linear regression model of cSFRP5 (continuous) versus TNM stage.

| Predictor | Comparison | Unadjusted |  | Adjusted by age and sex |  |
| --- | --- | --- | --- | --- | --- |
|  |  | Mean difference (95% CI) | p value | Mean difference (95% CI) | p value |
| TNM stage | I vs II | -9.76 (-12.42, -7.10) | **<0.0001** | -9.81 (-12.43, -7.20) | **<0.0001** |
|  | I vs III | -4.50 (-7.12, -1.89) | **0.0007** | -4.65 (-7.23, -2.07) | **0.0004** |
|  | I vs IV | -0.80 (-3.57, 1.97) | 0.5725 | -2.21 (-5.03, 0.60) | 0.1234 |
|  | II vs III | 5.26 (2.41, 8.10) | **0.0003** | 5.16 (2.36, 7.96) | **0.0003** |
|  | II vs IV | 8.96 (5.98, 11.95) | **<0.0001** | 7.60 (4.58, 10.61) | **<0.0001** |
|  | III vs IV | 5.10 (1.69, 8.55) | **0.0054** | 3.70 (0.76, 6.65) | **0.0138** |
| Age | Continuous |  |  | 1.69 (0.83, 2.56) | **<0.0001** |
| Sex | Female vs Male |  |  | 0.08 (-2.16, 2.32) | 0.9444 |

**Supplementary Table 6.** Linear regression model of cSFRP5 (continuous) versus patient groups.

| Predictor | Comparison | Unadjusted |  | Adjusted by age and sex |  |
| --- | --- | --- | --- | --- | --- |
|  |  | Mean difference (95% CI) | p value | Mean difference (95% CI) | p value |
| Patient group | H vs D | -11.97 (-15.17, -8.76) | **<0.0001** | -9.36 (-12.72, -6.00) | **<0.0001** |
|  | H vs P | -8.08 (-11.00, -5.15) | **<0.0001** | -4.97 (-8.12, -1.81) | **0.0020** |
|  | H vs CRC | -8.41 (-10.49, -6.33) | **<0.0001** | -5.07 (-7.54, -2.60) | **<0.0001** |
|  | D vs P | 3.89 (0.40, 7.37) | **0.0288** | 4.39 (0.93, 7.86) | **0.0130** |
|  | D vs CRC | 3.56 (0.74, 6.37) | **0.0132** | 4.29 (1.48, 7.10) | **0.0027** |
|  | P vs CRC | -0.33 (-2.82, 2.16) | 0.7949 | -0.10 (-2.56, 2.36) | 0.9342 |
| Age | Continuous |  |  | 1.35 (0.78, 1.92) | **<0.0001** |
| Sex | Female vs Male |  |  | -0.94 (-2.53, 0.66) | 0.2498 |

**Supplementary Table 7.** Univariate and multivariable analysis of overall survival for all CRC patients.

| Predictor | Univariate |  | Multivariable |  |
| --- | --- | --- | --- | --- |
|  | Hazard ratio (95% CI) | p value | Hazard ratio (95% CI) | p value |
| **cSFRP5\*** | 1.01 (0.99-1.02) | 0.4866 | 0.99 (0.98-1.02) | 0.9721 |
| **Age\*** | 1.03 (1.02-1.05) | **<0.001** | 1.06 (1.04-1.08) | **<0.001** |
| **Sex** |  |  |  |  |
| Male | 1 |  | 1 |  |
| Female | 0.86 (0.61-1.22) | 0.4066 | 1.06 (0.74-1.53) | 0.7331 |
| **TNM stage** |  |  |  |  |
| I | 1 |  | 1 |  |
| II | 1.33 (0.78-2.27) | 0.2894 | 1.24 (0.70-2.19) | 0.4526 |
| III | 1.91 (1.13-3.21) | **0.0151** | 1.69 (0.94-3.05) | 0.0943 |
| IV | 5.56 (3.55-8.72) | **<0.001** | 8.58 (5.16-14.25) | **<0.001** |
| **Differentiation** |  |  |  |  |
| Low (Grade 1-2) | 1 |  | 1 |  |
| High (Grade 3-4) | 1.23 (0.81-1.86) | 0.3286 | 1.15 (0.74-1.79) | 0.5442 |
| **Vascular or perineural invasion** |  |  |  |  |
| Negative | 1 |  | 1 |  |
| Positive | 2.27 (1.51-3.42) | **<0.001** | 1.59 (1.02-2.49) | **0.0402** |
| **Tumour site** |  |  |  |  |
| Colon | 1 |  | 1 |  |
| Rectum | 0.72 (0.49-1.07) | 0.1049 | 1.25 (0.81-1.91) | 0.3084 |

*\*Analysed as a continuous variable.*

